# Changes to the sebum lipidome upon COVID-19 infection observed via rapid sampling from the skin

**DOI:** 10.1101/2020.09.29.20203745

**Authors:** Matt Spick, Katie Longman, Cecile Frampas, Holly Lewis, Catia Costa, Deborah Dunn Walters, Alex Stewart, Mike Wilde, Danni Greener, George Evetts, Drupad Trivedi, Perdita Barran, Andy Pitt, Melanie Bailey

**Author notes:** **Corresponding Author:** Dr Melanie Bailey. **Sources of support:** The authors would like to acknowledge funding from the EPSRC Impact Acceleration Account for sample collection, as well as EPSRC Fellowship Funding EP/R031118/1. Mass Spectrometry was funded under EP/P001440/1.

## Abstract

The COVID-19 pandemic has led to an urgent and unprecedented demand for testing – both for diagnosis and prognosis. Here we explore the potential for using sebum, collected via swabbing of a patient’s skin, as a novel sampling matrix to fulfil these requirements. In this pilot study, sebum samples were collected from 67 hospitalised patients (30 PCR positive and 37 PCR negative). Lipidomics analysis was carried out using liquid chromatography mass spectrometry. Lipid levels were found to be depressed in COVID-19 positive participants, indicative of dyslipidemia. Partial Least Squares-Discriminant Analysis (PLS-DA) modelling showed promising separation of COVID-19 positive and negative participants when comorbidities and medication were controlled for, with sensitivity of 75% and specificity of 81% in stratified subsets. Given that sebum sampling is rapid and non-invasive, this work highlights the potential of this alternative matrix for testing for COVID-19.

## 1. Introduction

SARS-CoV-2, a novel coronavirus, was identified by the World Health Organization as originating in the Wuhan province of China in late 2019, ^1,2^ and causes <u>Co</u>rona <u>Vi</u>rus <u>Di</u>sease 20<u>19</u> (COVID-19). Mass testing has been identified by the World Health Organisation as a key weapon in the battle against COVID-19 to contain outbreaks and reduce hospitalisations. ^3^ Current approaches to testing require the detection of SARS-CoV-2 viral RNA collected from the upper respiratory tract via polymerase chain reaction (PCR). Whilst these types of tests are easily deployable and highly selective for the virus, they suffer from a significant proportion of false negative events; in addition, scarcity of reagents can be an issue for the scale of testing required. Furthermore, currently deployed approaches carry no prognostic information.

Approaches that measure the effect of the virus on the host (as opposed to direct measurement of the virus itself) may offer a complementary solution in clinical or mass testing settings; for example, one feasibility study has recently identified derangement of breath biochemistry in COVID-19 patients. ^4^ As the coronavirus requires lipids for reproduction, COVID-19 can be expected to disrupt the lipidome ^5^. Evidence of a dysregulated lipidome has been observed in patients with COVID-19 via analyses of blood plasma; ^6,7,8,9^ dysregulation of the skin would also be consistent with the ability of canines to differentiate COVID-19 positive and negative by smell. ^10^ Lipidomics therefore offers a promising route to better understanding of - and potentially diagnosis for - COVID-19. Sebum is a biofluid secreted by the sebaceous glands and is rich in lipids. A sample can be collected easily and non-invasively via a gentle swab of skin areas rich in sebum (for example the face, neck or back). Characteristic features have previously been identified from sebum for a limited number of illnesses such as Parkinson’s Disease and Type 1 Diabetes Mellitus. ^11,12,13^ In addition, whilst the mechanisms for the role of sebum in barrier function are not fully described, sebum lipids barrier function directly and also through commensal bacteria interactions; lipid dysregulation would have implications for skin health. ^14^ In this work, we explore differences in sebum lipid profiles for patients with and without COVID-19, with a view to exploring sebum’s future use as a non-invasive sampling medium for testing, as well as expanding the understanding of sebum as a sampling matrix.

In May 2020 several UK bodies announced their intention to pool resources and form the COVID-19 International Mass Spectrometry (MS) Coalition. ^15^ This consortium has the proximal goal of providing molecular level information on SARS-CoV-2 in infected humans, with the distal goal of understanding the impact of the novel coronavirus on metabolic pathways in order to better diagnose and treat cases of COVID-19 infection. This work took place as part of the COVID-19 MS Coalition and all data will be stored and fully accessible on the MS Coalition open repository.

## 2. Methods

### 2.1 Participant recruitment and ethics

Ethical approval for this project (IRAS project ID 155921) was obtained via the NHS Health Research Authority (REC reference: 14/LO/1221). The participants included in this study were recruited at NHS Frimley Park NHS Trust, totalling 67 participants. Collection of the samples was performed by researchers from the University of Surrey at Frimley Park NHS Foundation Trust hospitals. Participants were identified by clinical staff to ensure that they had the capacity to consent to the study, and were asked to sign an Informed Consent Form; those that did not have this capacity were not sampled. Consenting participants were categorised by the hospital as either “query COVID” (meaning there was clinical suspicion of COVID-19 infection) or “COVID positive” (meaning that a positive COVID test result had been recorded during their admission). All participants were provided with a Patient Information Sheet explaining the goals of the study.

### 2.2 Sample collection, inactivation and extraction

Patients were sampled immediately upon recruitment to the study. This meant that the range in time between symptom onset and sebum sampling ranged from 1 day to > 1 month, an inevitable consequence of collecting samples in a pandemic situation. Each participant was swabbed on the right side of the upper back, using 15 cm by 7.5 cm gauzes that had each been folded twice to create a four-ply swab. The surface area of sampling was approximately 5 cm x 5 cm, pressure was applied uniformly whilst moving the swab across the upper back for ten seconds. The gauzes were placed into Sterilin polystyrene 30 mL universal containers.

Samples were transferred from the hospital to the University of Surrey by courier within 4 hours of collection, whereupon the samples were then quarantined at room temperature for seven days to allow for virus inactivation. Finally, the vials were transferred to minus 80°C storage until required. Alongside sebum collection, metadata for all participants was also collected covering *inter alia* sex, age, comorbidities (based on whether the participant was receiving treatment), the results and dates of COVID PCR (polymerase chain reaction) tests, bilateral chest X-Ray changes, smoking status, and whether the participant presented with clinical symptoms of COVID-19. Values for lymphocytes, CRP and eosinophils were also taken - here the most extreme values during the hospital admission period were recorded. These were not collected concomitantly with the sebum samples.

The extraction, storage and reconstitution of the obtained samples followed Sinclair E, Trivedi D, Sarkar D, *et al*. ^16^ Samples were analysed over a period of five days. Each day consisted of a run incorporating solvent blank injections (n=5), pooled QC injections (n=3), followed by 16 participant samples (triplicate injections of each) with a single pooled QC injection every six injections. Each day’s run was completed with pooled QC injections (n=2) and solvent blanks (n=3). A triplicate injection of a field blank was also obtained.

### 2.3 Instrumentation and software

Analysis of samples was carried out using a Dionex Ultimate 3000 HPLC module equipped with a binary solvent manager, column compartment and autosampler, coupled to a Orbitrap Q-Exactive Plus mass spectrometer (Thermo Fisher Scientific, UK) at the University of Surrey’s Ion Beam Centre. Chromatographic separation was performed on a Waters ACQUITY UPLC BEH C18 column (1·7 µm, 2·1 mm x 100 mm) operated at 55 °C with a flow rate of 0·3 ml min^-1^.

The mobile phases were as follows: mobile phase A was acetonitrile:water (v/v 60:40) with 0·1% formic acid, whilst mobile phase B was 2-propanol:acetonitrile (v/v, 90:10) with 0·1% formic acid (v/v). An injection volume of 5 µL was used. The initial solvent mixture was 40% B, increasing to 50% B over 1 minute, then to 69% B at 3·6 minutes, with a final ramp to 88% B at 12 minutes. The gradient was reduced back to 40% B and held for 2 minutes to allow for column equilibration. Analysis on the Q-Exactive Plus mass spectrometer was performed in split-scan mode with an overall scan range of 150 *m/z* to 2 000 *m/z*, and 5 ppm mass accuracy. Split scan was chosen to extend the *m/z* range from 150 to 2 000 *m/z* whilst maximising the number of features identified ^17,18^. MS/MS validation of features was carried out on Pooled QC samples using data dependent acquisition mode. Operating conditions are summarised in Table S1 (Supplementary Information).

### 2.4 Materials and chemicals

The materials and solvents utilised in this study were as follows: gauze swabs (Reliance Medical, UK), 30 mL Sterilin™ tubes (Thermo Scientific, UK), 10 mL syringes (Becton Dickinson, Spain), 2 mL microcentrifuge tubes (Eppendorf, UK), 0.2 µm syringe filters (Corning Incorporated, USA), 200 µL micropipette tips (Starlab, UK) and Qsert™ clear glass insert LC vials (Supelco, UK). Optima™ (LC-MS) grade methanol was used as an extraction solvent, and Optima™ (LC-MS) grade methanol, ethanol, acetonitrile and 2-propanol were used to prepare injection solvents and mobile phases. Formic acid was added to the mobile phase solvents at 0.1% (v/v). Solvents were purchased from Fisher Scientific, UK.

### 2.5 Data processing

LC-MS outputs (.raw files) were pre-processed for alignment, normalisation and peak identification using Progenesis QI (Non-Linear Dynamics, Waters, Wilmslow, UK), a platform-independent small molecule discovery analysis software for LC-MS data. Peak picking (mass tolerance ±5 ppm), alignment (RT window ±15 s) and area normalisation was carried out with reference to the pooled QC samples. Features identified in MS were initially annotated using accurate mass match with Lipid Blast in Progenesis QI, whilst validation was performed using data dependent MS/MS analysis using LipidSearch (Thermo Fisher Scientific, UK) and Compound Discoverer (Thermo Fisher Scientific, UK). This process yielded an initial peak table with 14,160 features. All those features with a coefficient of variation across all pooled QCs above 20% were removed, as were those that were not present in at least 90% of pooled QC injections. These features were then field blank adjusted: all those features with a signal to noise ratio below 3x were also rejected. The remaining set of 998 features were deemed to be robust, reproducible and suitably distinct from those found in the field blank.

Inclusion criteria were also applied to participant data, requiring both full completion of metadata and also agreement between the result of the PCR COVID-19 test (Y/N) and the clinical diagnosis for COVID-19 (Y/N). Whilst these inclusion criteria reduced the total number of participants from n=87 to n=67, this was considered worthwhile given the potential for misdiagnosis to confound the development of statistical models.

### 2.6 Statistical Analysis

Data processing and analysis of the pareto-scaled peak:area matrix was conducted through a combination of the R package mixOmics, ^19^ supplemented by user-written scripts in the statistical programming language R. ^20^ PLS-DA was used for classification and prediction of data. Separation and classification was based on mahalabonis distance between observations. Leave-one-out cross-validation was used for PLS-DA model validation to test accuracy, sensitivity and specificity; variable importance in projection (VIP) scores were used to assess feature significance.

## 3. RESULTS

### 3.1 Population metadata overview

The study population analysed in this work included 67 participants, comprising 30 participants presenting with COVID-19 clinical symptoms (and an associated positive COVID-19 RT-PCR test) and 37 participants presenting without. A summary of the metadata is shown in Table 1.

**Table 1.** Summary of clinical characteristics by participant cohort

| Parameters | Negative for COVID-19 | Positive for COVID-19 |
| --- | --- | --- |
| n | 37 | 30 |
| Age (mean, standard deviation; years) | 65.0 ± 19.3 | 64.7 ± 19.4 |
| Male / Female (n) | 18 / 19 | 17 / 13 |
| Treated for Hypertension (n) | 17 | 10 |
| Treated for High Cholesterol (n) | 10 | 5 |
| Treated for Type 2 Diabetes Mellitus (n) | 12 | 7 |
| Treated for Ischemic Heart Disease (n) | 7 | 4 |
| Current Smoker (n) | 1 | 0 |
| Ex-Smoker (n) | 10 | 4 |
| Medical Acute Dependency admission (n) | 4 | 11 |
| Intensive Care Unit admission (n) | 0 | 5 |
| Survived Admission (n) | 35 | 27 |
| Lymphocytes (mean, standard deviation; cells / μL) | 1.0 ± 0.5 | 0.6 ± 0.3 |
| C-Reactive Protein (mean, standard deviation; mg / L) | 132.1 ± 95.7 | 181.3 ± 117.2 |
| Eosinophils (mean, standard deviation; 100 / μL) | 0.3 ± 0.4 | 0.2 ± 0.4 |
| Bilateral Chest X-Ray changes (n) | 2 | 21 |
| Continuous Positive Airway Pressure (n) | 2 | 10 |
| O<sub>2</sub> required (n) | 10 | 20 |

There were more male participants in the COVID-19 positive group (M:F ratio of 0·57) compared to the participant population overall (M:F ratio of 0·52); given recruitment took place in a hospital environment, this may reflect increased severity amongst males. ^21^ Age distributions for COVID-19 positive and negative cohorts were almost identical (mean age of 64·7 years and 65·0 years respectively). Comorbidities are associated with both hospitalisation and more severe outcomes for COVID-19 infection, but will also alter the metabolome of participants, representing both a causative and confounding factor. The impact on classification accuracy of these comorbidities was tested by stratifying participant data by comorbidity to see if separation improved; this process is described in the following sections. In this pilot study, comorbidities were less well represented in the cohort of COVID-19 positive participants than in the cohort of COVID-19 negative participants.

Levels of C-Reactive Protein (CRP) were significantly higher for COVID-19 participants, whilst lymphocyte and eosinophils levels were lower. A two-tailed Mann Whitney U test on the CRP indicator provided a p-value of 0·031, and on the lymphocytes a p-value of 0·004. Effect sizes (calculated by Cohen’s D) were 0·56 and 0·85 respectively. COVID-19 positive participants were also more likely to present with bilateral chest X-ray changes (21 out of 30 COVID-19 positive patients, versus 2 out of 37 COVID-19 negative patients). COVID-19 positive participants experienced higher rates of requiring oxygen / CPAP, higher rates of escalation, and lower survival rates. These observations were in agreement with literature descriptions of COVID-19 symptoms and progression. ^22^

### 3.2 Overview of features identified by Liquid Chromatography Mass Spectrometry (LC-MS)

998 features were identified reproducibly by LC-MS (present in greater than 90% of pooled QC LC-MS injections, coefficient of variation below 20% across pooled QCs, signal to noise ratio greater than three) and these formed the basis of the analysis in this work. Differences between COVID-19 positive and negative participants were observed across a range of lipids and metabolites, with the most consistent difference seen in reduced lipid levels, especially triglycerides (Figure 1).

**Figure 1:**
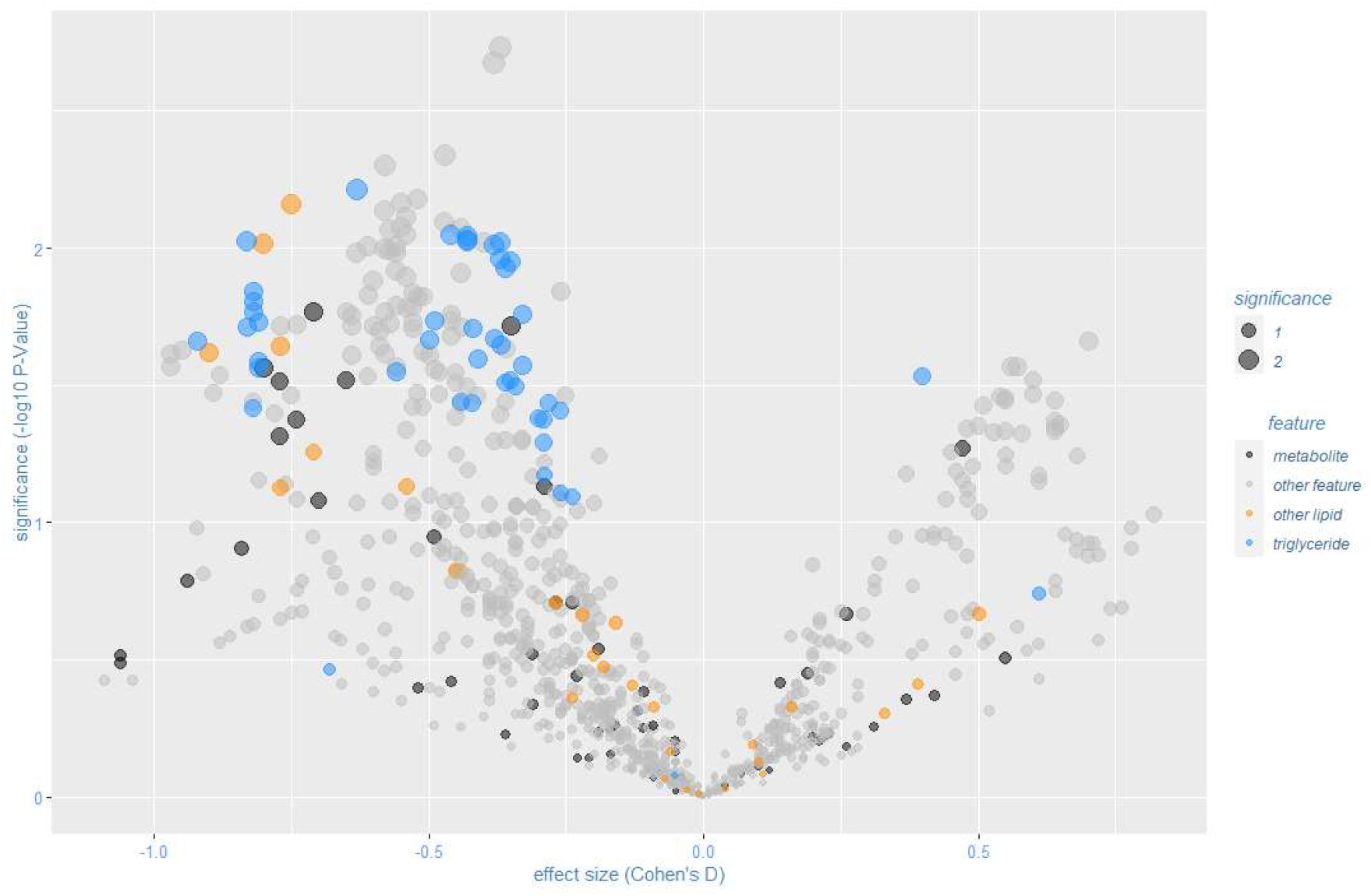
Volcano plot of features for COVID-19 positive (n=30) versus negative (n=37), labelled features validated by MS/MS, points scaled to significance

Aggregate levels of triglycerides identified by MS/MS were depressed for COVID-19 positive participants, and also for ceramides, albeit fewer lipids of the latter class were identified and validated. The distributions of the natural log of aggregated lipid ion counts by class were not characterised as normal by Shapiro-Wilk normality tests. ^23^ Two-tailed Mann-Whitney U-tests were performed to test the significance of aggregate levels of these lipid classes. These resulted in p-values of 0·022 and 0·015 for triglycerides and ceramides respectively, with effect sizes (calculated by Cohen’s D) of 0·44 and 0·57, indicative of medium effect size. These results are suggestive of dyslipidemia within the stratum corneum due to COVID-19. The alteration in levels of triglycerides between positive and negative cohorts is comparable to that for CRP or for lymphocytes as indicators of COVID-19 status (Figure 2).

**Figure 2:**
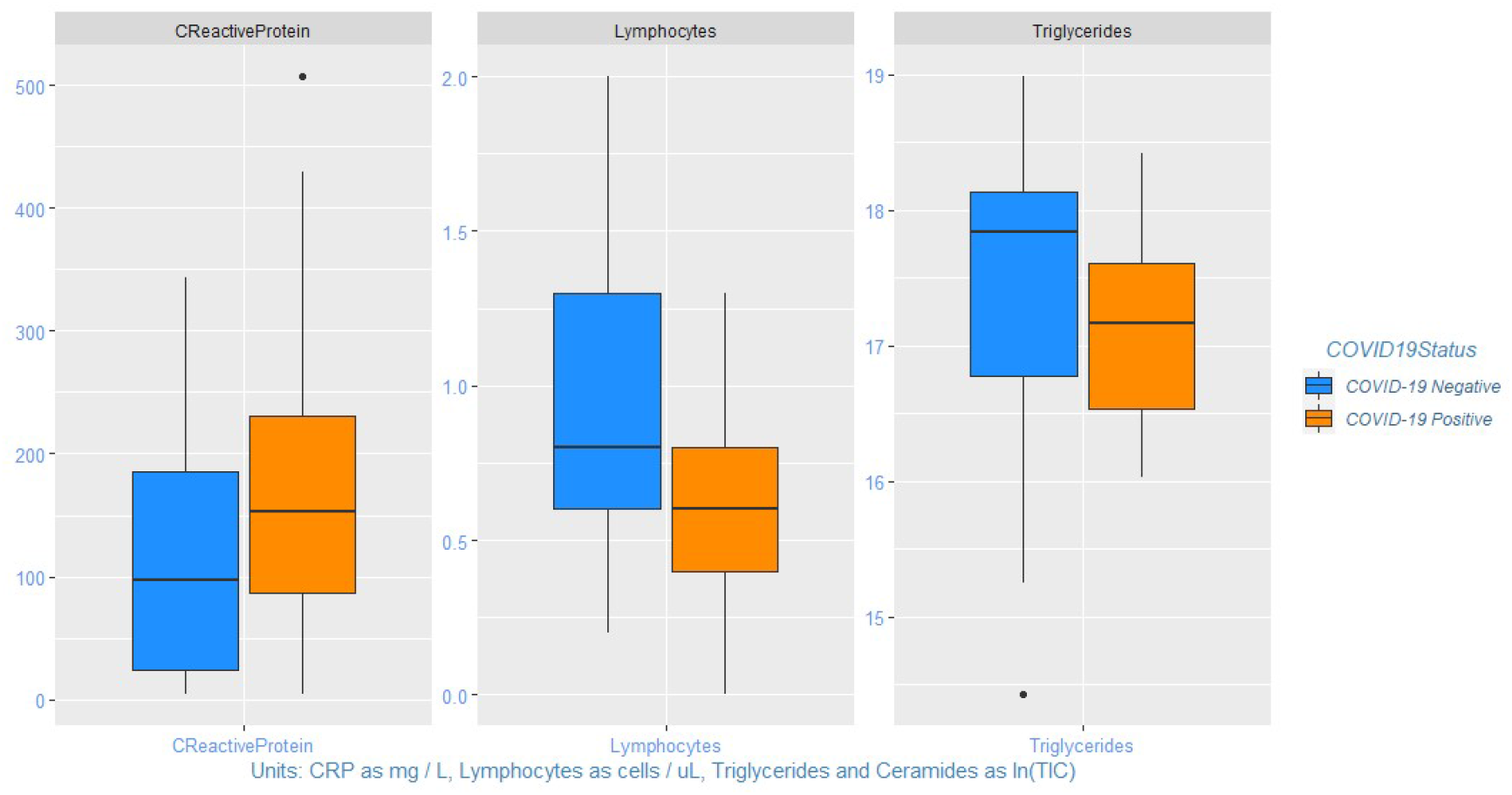
Boxplots of diagnostic indicators versus triglyceride levels

Other work has found evidence of dyslipidemia in plasma from COVID-19 positive patients, ^7,6,9^ although evidence of whether upregulation or downregulation is dominant for these lipid classes is mixed. Plasma triglyceride (TAG) levels have been found to be elevated in blood plasma for mild cases of COVID-19, but TAG levels in plasma may also decline as the severity of COVID-19 increased. ^24^

It should be remembered, however, that the primary role of skin is barrier function, and lipid expression in the stratum corneum depends on *de novo* lipogenesis – in fact nonskin sources such as plasma provide only a minor contribution to sebum lipids, ^25^ which limits the relevance of broader pathway analysis to this biofluid. To the extent that the virus sequesters lipids for its own reproduction, it is possible that this causes deficiency in the expression of sebum lipids.

### 3.3 Population-level clustering analyses

No clustering was identifiable at the total population level by principal component analysis (PCA), i.e. by unsupervised analysis. Partial least squares discriminant analysis (PLS-DA) performed on the same data set revealed limited separation (Figure 3), with the area under the receiver operating curve (AUROC) over two components of 0.88. AUROC can be inflated when only used on a single training data set, and so a confusion matrix was constructed using a leave-one-out approach. Validating accuracy in this way (Table 2) showed sensitivity of just 57% and specificity of 68%. Given the wide range of comorbidities, this is not unexpected.

**Table 2.** Confusion matrix for COVID-19 positive versus negative (all participants)

| All Participants (n=67) | True COVID-19 Positive (n=30) | True COVID-19 Negative (n=37) |
| --- | --- | --- |
| Predicted COVID-19 Positive (% , n) | 57% (17) | 32% (12) |
| Predicted COVID-19 Negative (% , n) | 43% (13) | 68% (25) |

**Figure 3:**
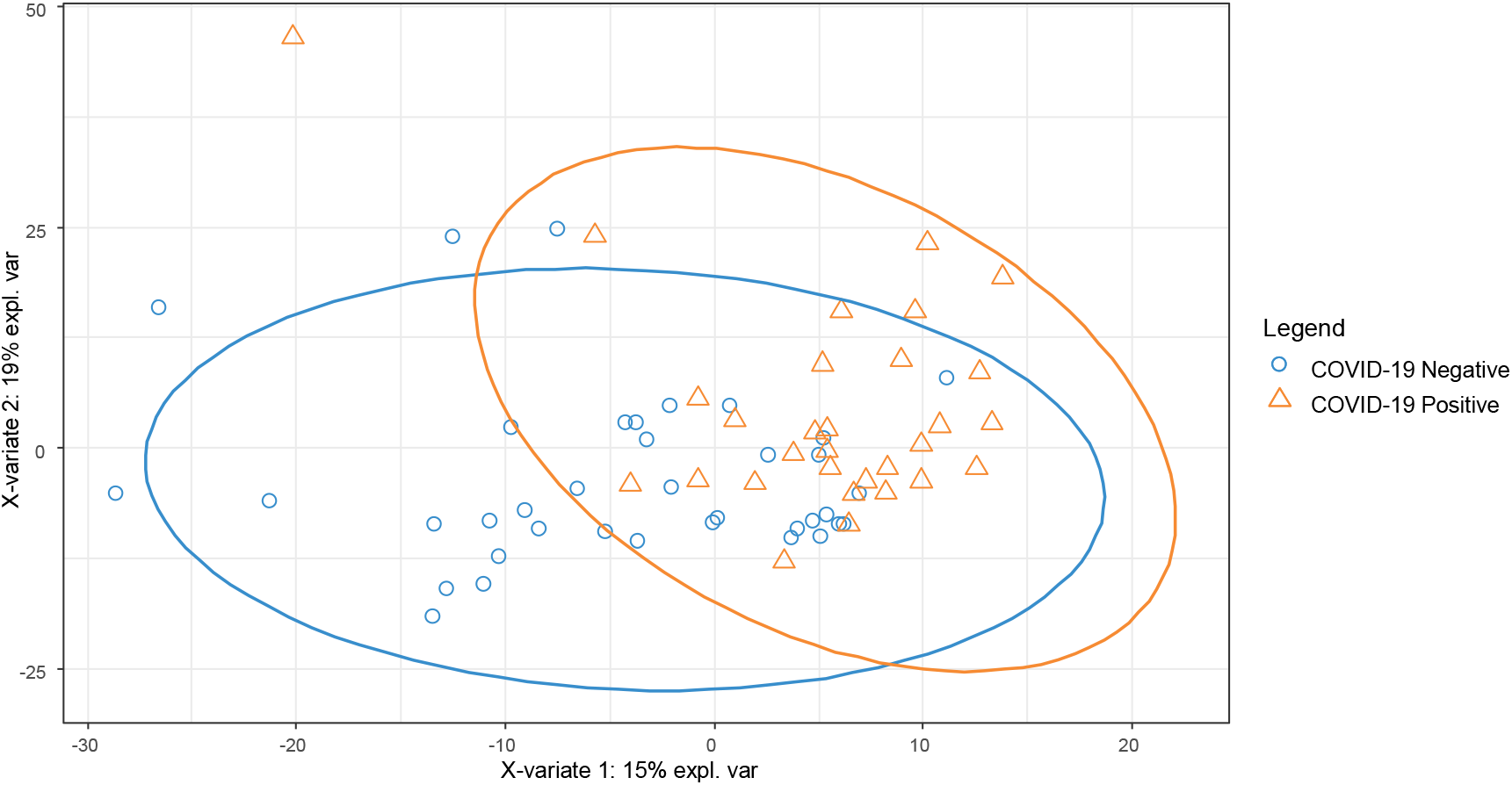
PLS-DA plot for 67 participants, classified by COVID-19 positive / negative

### 3.4 Investigation of confounding factors

To test the impact of age and diagnostic indicators (CRP, lymphocytes and eosinophils), these variables were pareto-scaled and included in the matrix for PLS-DA modelling. Variable importance in projection (VIP) scores for lymphocytes, CRP, and eosinophils were 2·47, 1·77 and 0.72 respectively, ranking 1, 90 and 465 out of 1,002 total features. As a single feature, depressed lymphocyte levels show high correlation with COVID-19 positive status, consistent with lymphocyte count being both a diagnostic and prognostic biomarker. ^26^ Age as a vector had a VIP score of just 0·05 (ranking 958 out of 1,002 total features), indicating that age is a smaller influencer of stratum corneum lipids than other factors.

Overall, PLS-DA separation improved by the addition of lymphocyte and CRP indicators, with slight model accuracy increases when these two variables were included in the feature matrix (from 62% to 64% accuracy for the overall population, for example). Given that this work focuses on sebum sampling, however, in the analyses that follow only features obtained from sebum are included, i.e. information from other diagnostic indicators is excluded from classification models.

To test whether separation based on sebum alone would improve in smaller / more homogenous groups, separate PLS-DA models were built for each split of the population by comorbidity. If model performance improved (measured by predictive power - Q2Y - and sensitivity and specificity via leave-one-out cross validation) then this could indicate that sebum lipid profiling would perform better if models were constructed based on stratified and matched datasets. Table shows the results for these metrics across the different modelled subsets.

Separation generally improved as the data were grouped more finely and modelled predictive power improved. Based on a weighted mean average for these subsets, sensitivity improved to 75% and specificity improved to 81%. For example, PLS-DA modelling of the subset of participants under medication for hypertension (Figure 4) showed both good separation and better sensitivity and specificity (Table 4). These data suggest that comorbidities are confounders in skin lipidomics.

**Table 3:** Summary of model parameters for different population subsets

| Population | Category 1 (n) | Category 2 (n) | Accuracy | Sensitivity | Specificity |
| --- | --- | --- | --- | --- | --- |
| All participants | COVID-19 Positive (30) | COVID-19 Negative (37) | 62% | 57% | 68% |
| Male | COVID-19 Positive (17) | COVID-19 Negative (18) | 66% | 65% | 67% |
| Female | COVID-19 Positive (13) | COVID-19 Negative (19) | 59% | 54% | 63% |
| Type 2 Diabetes Mellitus | COVID-19 Positive (7) | COVID-19 Negative (12) | 73% | 71% | 75% |
| High Cholesterol | COVID-19 Positive (5) | COVID-19 Negative (10) | 90% | 100% | 80% |
| Hypertension | COVID-19 Positive (10) | COVID-19 Negative (17) | 81% | 80% | 82% |
| IHD | COVID-19 Positive (4) | COVID-19 Negative (7) | 68% | 50% | 86% |
| Statins | COVID-19 Positive (11) | COVID-19 Negative (21) | 63% | 55% | 90% |

**Table 4:** Confusion matrix for COVID-19 positive versus negative (participants with hypertension)

| Hypertension (n=27) | True COVID-19 Positive (n=10) | True COVID-19 Negative (n=17) |
| --- | --- | --- |
| Predicted COVID-19 Positive (% , n) | 80% (8) | 18% (3) |
| Predicted COVID-19 Negative (% , n) | 20% (2) | 82% (14) |

**Figure 4:**
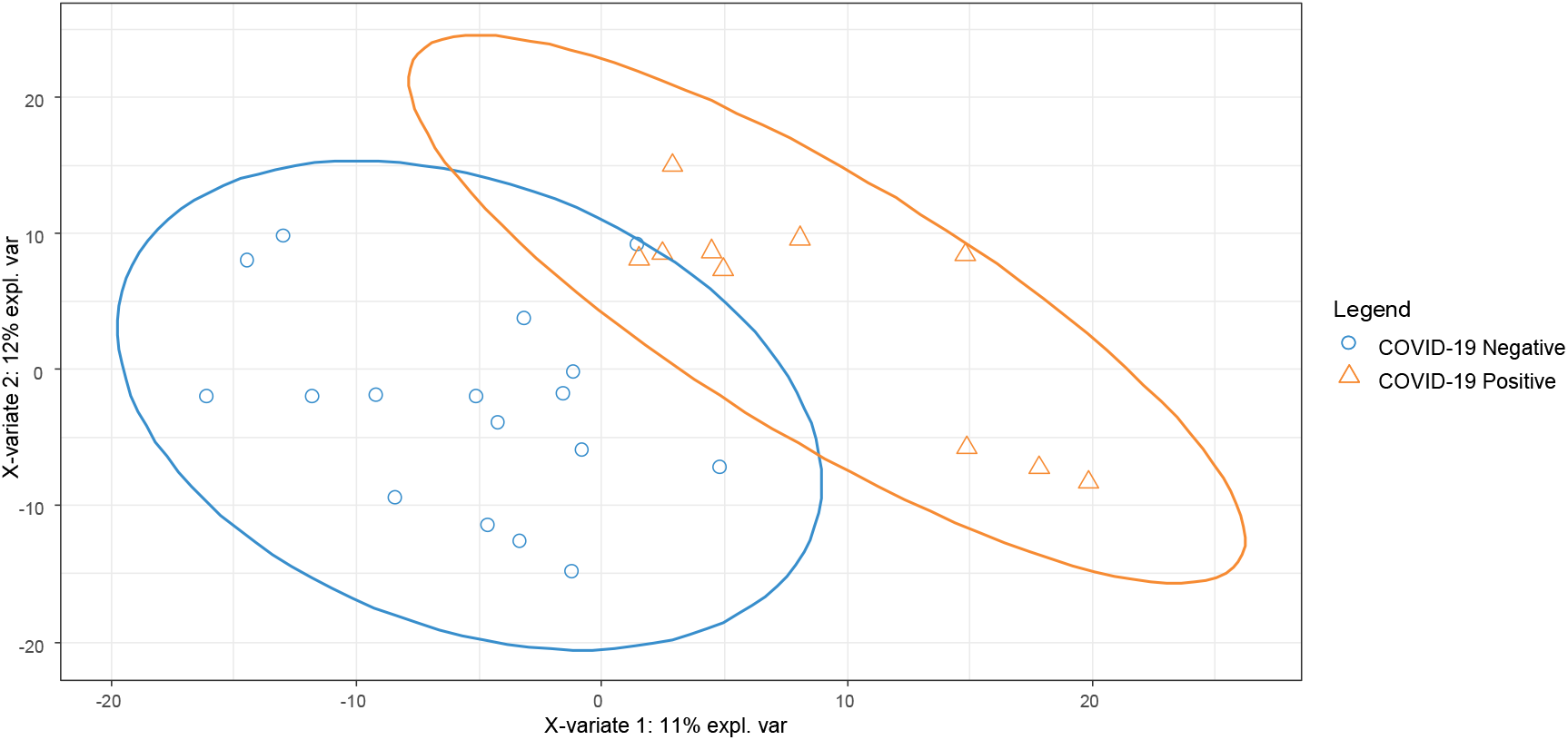
PLS-DA plot for 15 participants with hypertension, COVID-19 positive / negative

Similarly, PLS-DA modelling of the subset of participants under medication for high cholesterol showed good separation (Figure S1, Supplementary Information), with sensitivity of 100% and specificity of 80%. This subgroup was treated with lipid-lowering agents, specifically statins. The subgroup comprising participants undergoing treatment for ischemic heart disease (IHD) also showed much better separation (Figure S2, Supporting Material), with better overall accuracy, with sensitivity and specificity of 50% and 86% respectively. This subgroup received varied medication, but participants presenting with IHD were also being prescribed statins. Finally, the subset of participants under medication for T2DM (Figure 4) also showed both good separation and better sensitivity and specificity (of 71% and 75% respectively). This subgroup was typically being treated with oral hypoglycaemics, for example metformin, in some cases with insulin and in some instances with diet control only.

Model performance (Figure S4, Supporting Material) also improved versus the base population for a stratified dataset based on those participants taking statins (sensitivity of 55% and specificity of 90%). Given that statins control cholesterol and lipid levels, this may have provided a more similar “baseline” against which to measure perturbance in the lipidome by COVID-19; patients taking statins which included both participants treated for high cholesterol and also participants with poor diabetic control or history of ischaemic heart disease, where statins are routinely added prophylactically to improve long-term outcomes.

Looking across the models, there was commonality in the features identified as significant in differentiating between COVID-19 positive and negative. Many features featured in all subsets with VIP scores above 2 (dark grey in Figure 5), but others did not, a possible indicator of overfitting due to the smaller groups when stratified. Where overlap does occur between the features, this may reflect the natural overlap between the subset populations, for example the subsets of participants presenting with ischaemic heart disease and with high cholesterol are largely subsets of the participants receiving treatment by statins.

**Figure 5:**
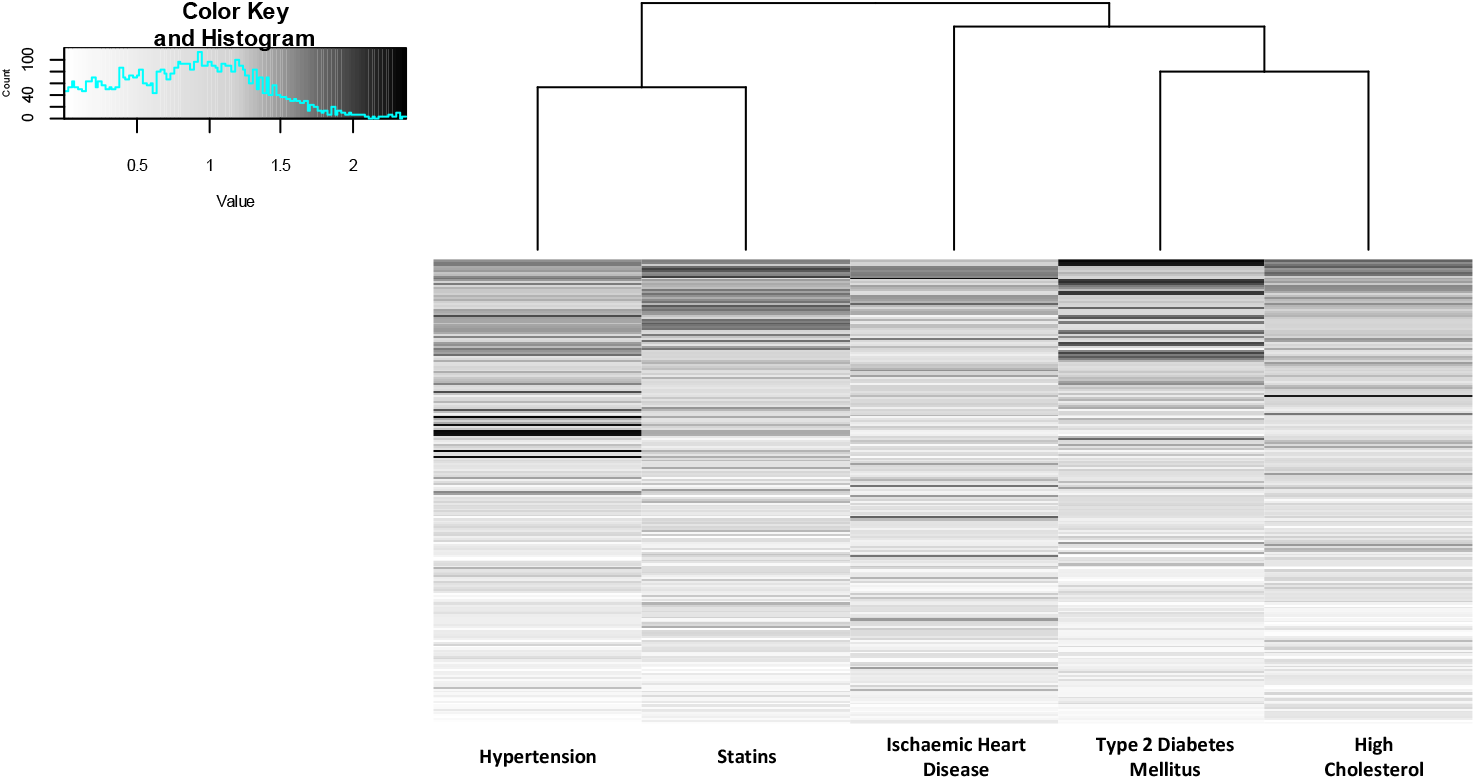
Heat map of VIP scores ranked by commonality to different subgroup PLS-DA models

Of the features with the highest common VIP scores, the two highest were triglycerides - TG(16.1/21.0/22.6) and TG(17.0/22.1/22.6) – as were eight of the highest twenty, consistent with previous observations that dysregulation of lipids, especially triglycerides, is a distinguishing feature of COVID-19’s impact on the skin.

## 4. Discussion

At the aggregate level, analysis of the metadata for the participants in this study illustrates the challenges involved in constructing a well-designed sample set during a pandemic. Age ranges of participants were large, and a wide range of comorbidities were present, leading to many confounding factors. Definitive separation has not proved possible in this pilot study, given that too few datapoints were available to rigorously stratify by medication or by comorbidity. Nonetheless, at the aggregate level, participants with a positive clinical COVID-19 diagnosis present with depressed lipid levels (triglycerides and ceramides in particular), with the possibility of reduced barrier function and skin health. Furthermore, these findings suggest that better stratification of participants could yield a clearer separation of positive and negative COVID-19 participants by their lipidomic profile. The overall accuracy in the stratified groups of 79% is comparable to that recently reported using breath biochemistry of 81%, ^4^ albeit overfitting is a risk in any pilot study with small *n*. This risk can only be reduced through both a larger training set of data and subsequently testing the models on future validation sets, made possible through cohesive efforts such as the work of the MS Coalition.

Another point to note is a possible lack of confounders in the participant population from seasonal respiratory viruses. Whilst the COVID-negative patients included patients with respiratory illnesses (e.g. COPD, asthma) and COVID-like symptoms, samples were collected between May and July, when the incidence of respiratory viruses is generally low. Both the common cold and influenza have some symptoms overlap with COVID-19 and may possibly lead to alterations to lipid metabolism that could interfere with the identification of features related to COVID-19 infection. Such viruses within the UK are more prevalent in autumn and winter. ^27^ Whilst it seems unlikely that seasonal respiratory viruses were a major confounding factor in this work, this is a factor that will need to be taken into account in future studies, and may also allow the opportunity to test sebum’s selectivity and specificity with regard to other respiratory viruses.

In conclusion, we provide evidence that COVID-19 infection leads to dyslipidemia in the stratum corneum. We further find that the sebum profiles of COVID positive and negative patients can be separated using the multivariate analysis method PLS-DA, with the separation improving when the patients are segmented in accordance with certain comorbidities. Given that sebum samples can be provided quickly and painlessly, we conclude that sebum is worthy of future consideration for clinical sampling for COVID-19 infection.

## Data Availability

This work took place as part of the COVID-19 MS Coalition and all data will be stored and fully accessible on the MS Coalition open repository.

## Data sharing statement

Participant metadata data with identifiers, alongside mass spectrometry .RAW files will be made available on the Mass Spectrometry Coalition website upon publication of this study. The analytical protocols used as well as sample and participant data will be openly available for all researchers to access. The website URL is https://covid19-msc.org/

## Author’s contributions

MPS was responsible for statistical analysis and authorship of the manuscript. MPS and MW extracted the samples, with MW providing additional advice on multivariate analysis. KL, CF and AS collected all samples used in this work; AS and DDW obtained ethical approval. GE and DG facilitated access to participants and collected participant metadata. PB, DT and AP advised on sampling protocol and on mass spectrometry acquisition and protocols. CC and HL assisted with mass spectrometry method development. MB obtained funding for the study, and was responsible for supervision of the research team.

## Acknowledgements

The authors acknowledge Samiksha Ghimire from Groningen Medical School for translation of participant information sheets and consent forms into Nepalese. The authors acknowledge Amanda Souza and Ioanna Ntai of Thermo Fisher, as well as Holly Lewis, Mason Malloy, Patrick Sears and Janella de Jesus of the University of Surrey, for their help with method development. We are grateful to Thanuja Weerasinge (Jay), Manjula Meda, Chris Orchard and Joanne Zamani of Frimley Park NHS Foundation Trust for their help with ethics approvals and access to hospital patients.

## Supplementary Information

**Table S1:**
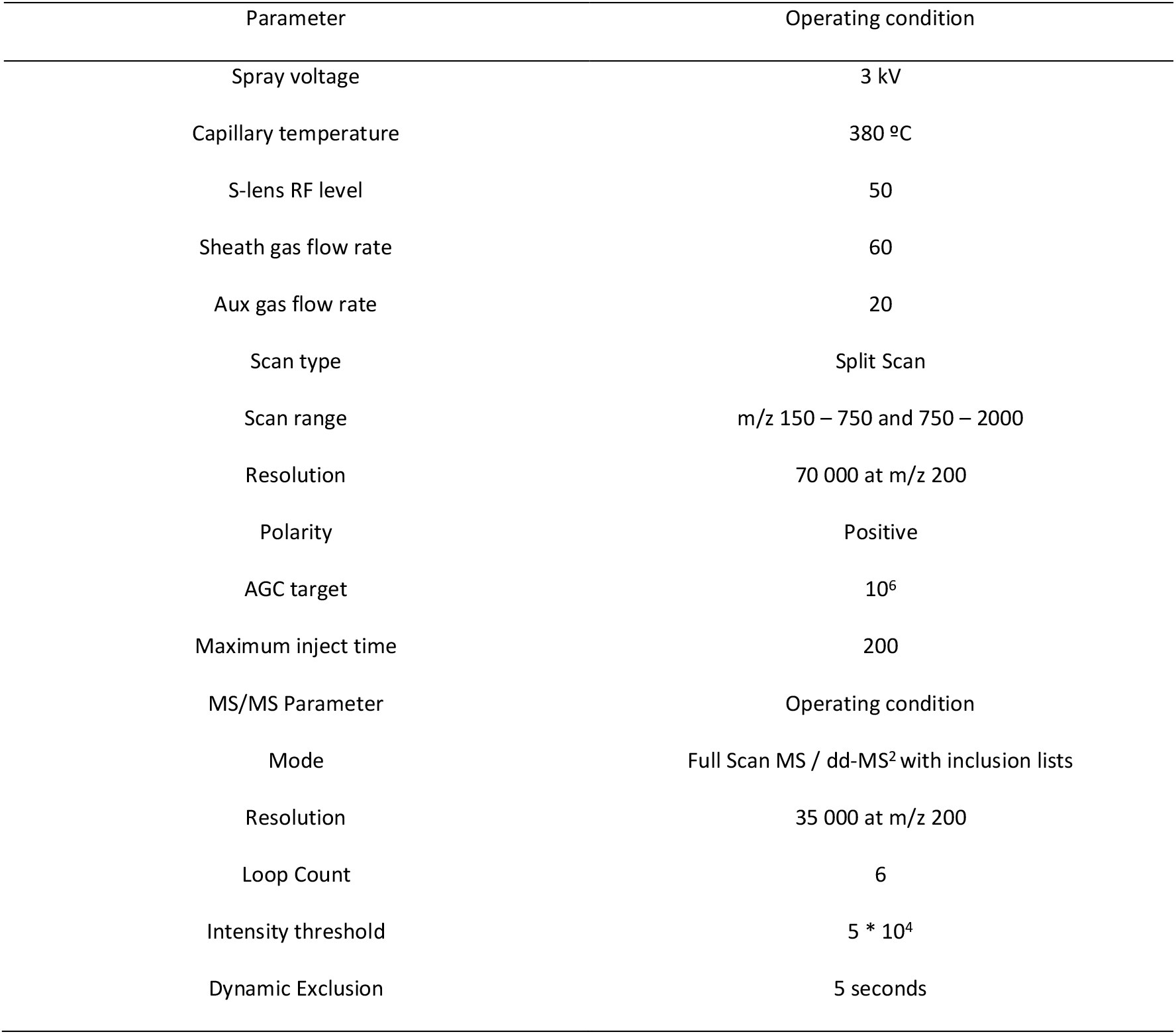
Operating conditions of the mass spectrometer used in this research

| Parameter | Operating condition |
| --- | --- |
| Spray voltage | 3 kV |
| Capillary temperature | 380 °C |
| S-lens RF level | 50 |
| Sheath gas flow rate | 60 |
| Aux gas flow rate | 20 |
| Scan type | Split Scan |
| Scan range | m/z 150 – 750 and 750 – 2000 |
| Resolution | 70 000 at m/z 200 |
| Polarity | Positive |
| AGC target | 10<sup>6</sup> |
| Maximum inject time | 200 |
| MS/MS Parameter | Operating condition |
| Mode | Full Scan MS / dd-MS<sup>2</sup> with inclusion lists |
| Resolution | 35 000 at m/z 200 |
| Loop Count | 6 |
| Intensity threshold | 5 * 10<sup>4</sup> |
| Dynamic Exclusion | 5 seconds |

**Table S2:** Confusion matrix for COVID-19 positive versus negative (participants with high cholesterol)

| High Cholesterol (n=15) | True COVID-19 Positive (n=5) | True COVID-19 Negative (n=10) |
| --- | --- | --- |
| Predicted COVID-19 Positive (% , n) | 100% (5) | 20% (2) |
| Predicted COVID-19 Negative (% , n) | 0% (0) | 80% (8) |

**Figure S1:**
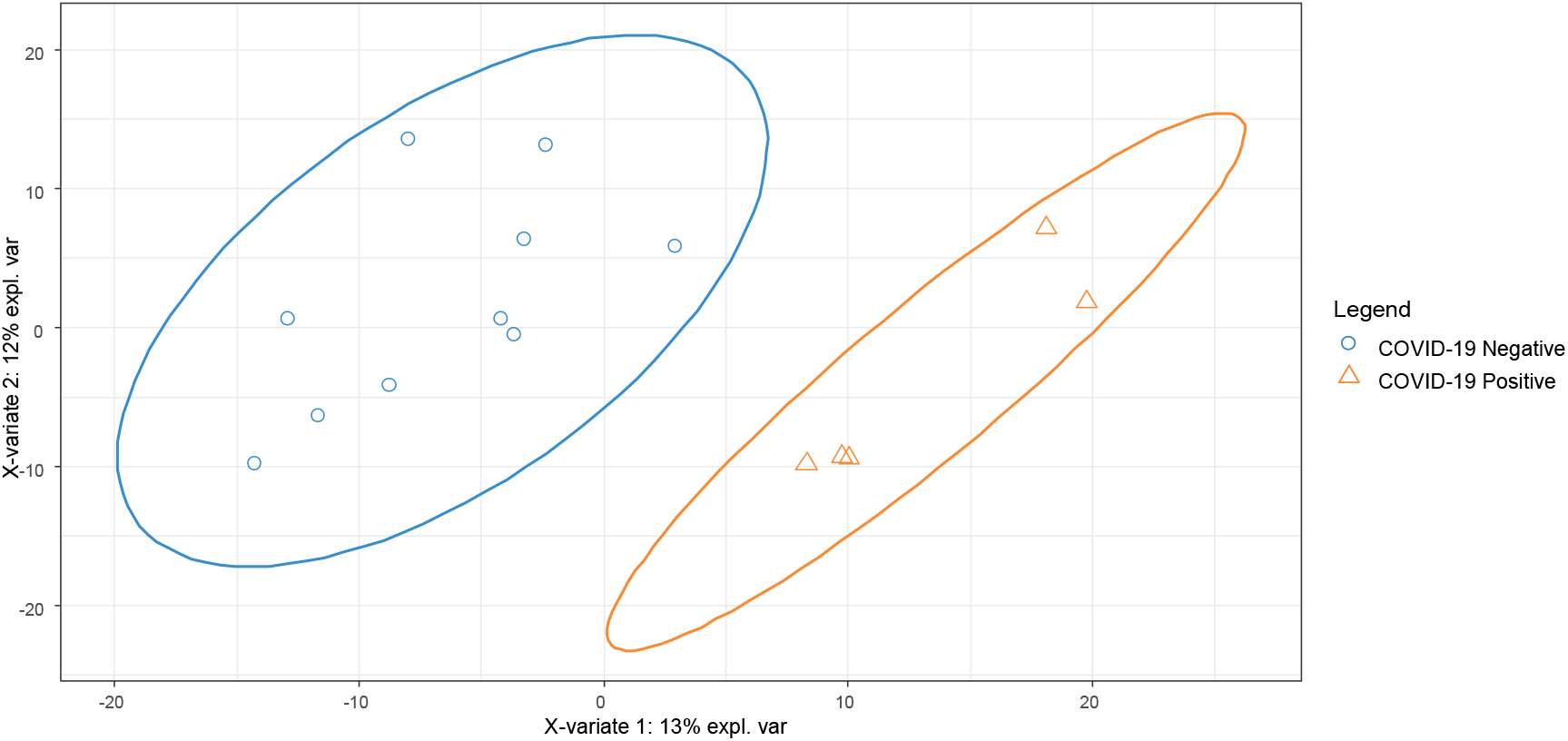
PLS-DA plot for 19 participants treated for high cholesterol, by COVID-19 positive / negative

**Table S3.** Confusion matrix for COVID-19 positive versus negative (participants with IHD)

| Participants with IHD (n=11) | True COVID-19 Positive (n=4) | True COVID-19 Negative (n=7) |
| --- | --- | --- |
| Predicted COVID-19 Positive (% , n) | 50% (2) | 14% (1) |
| Predicted COVID-19 Negative (% , n) | 50% (2) | 86% (6) |

**Figure S2:**
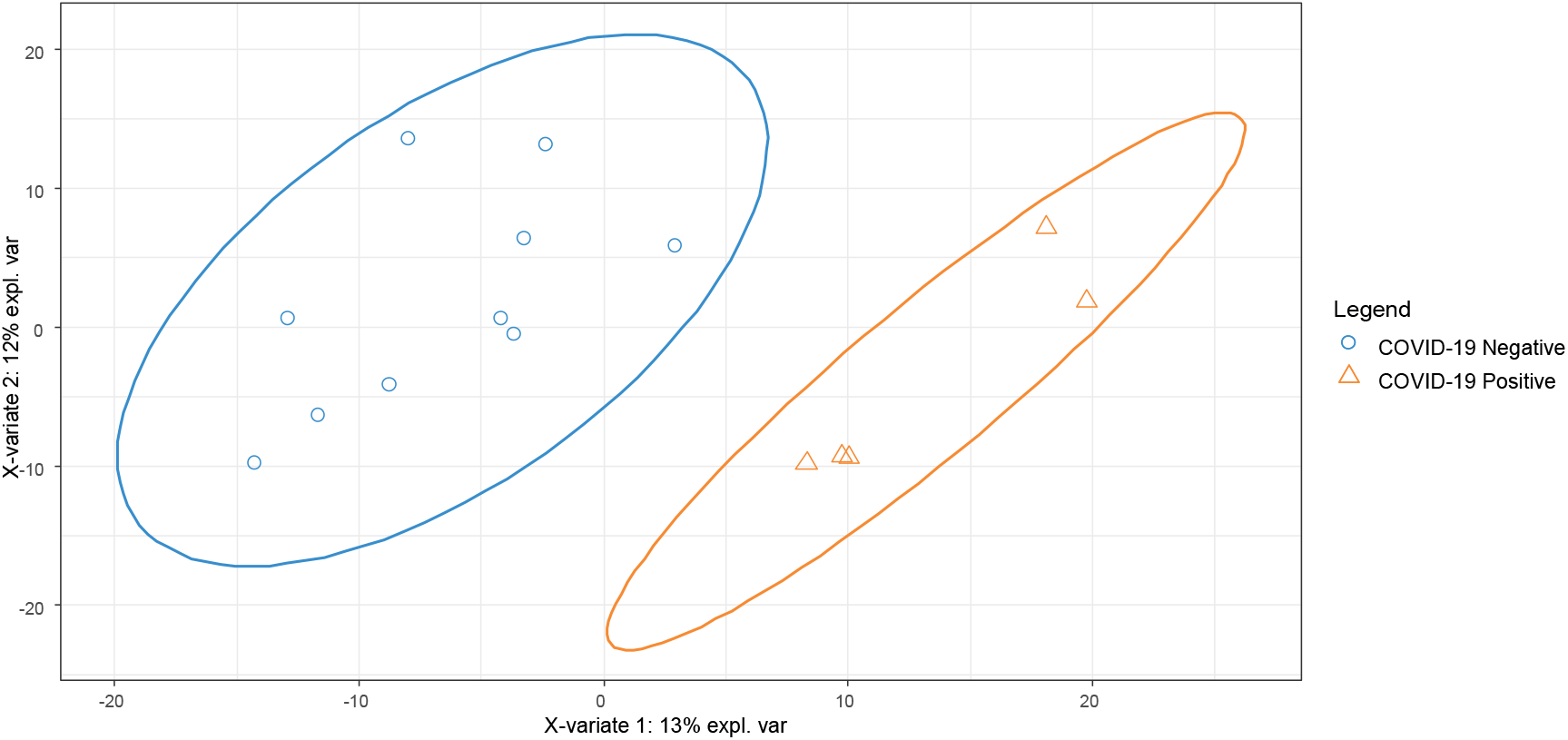
PLS-DA plot for 11 participants treated for IHD, by COVID-19 positive / negative

**Table S4.**
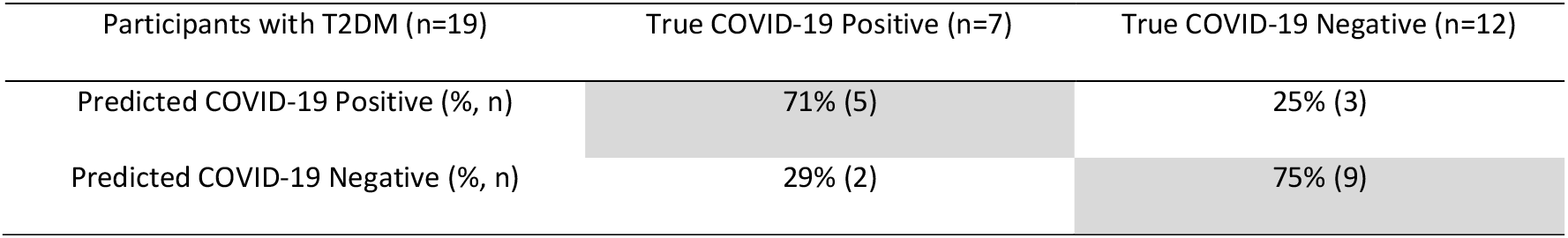
Confusion matrix for COVID-19 positive versus negative (participants with T2DM)

**Figure S3:**
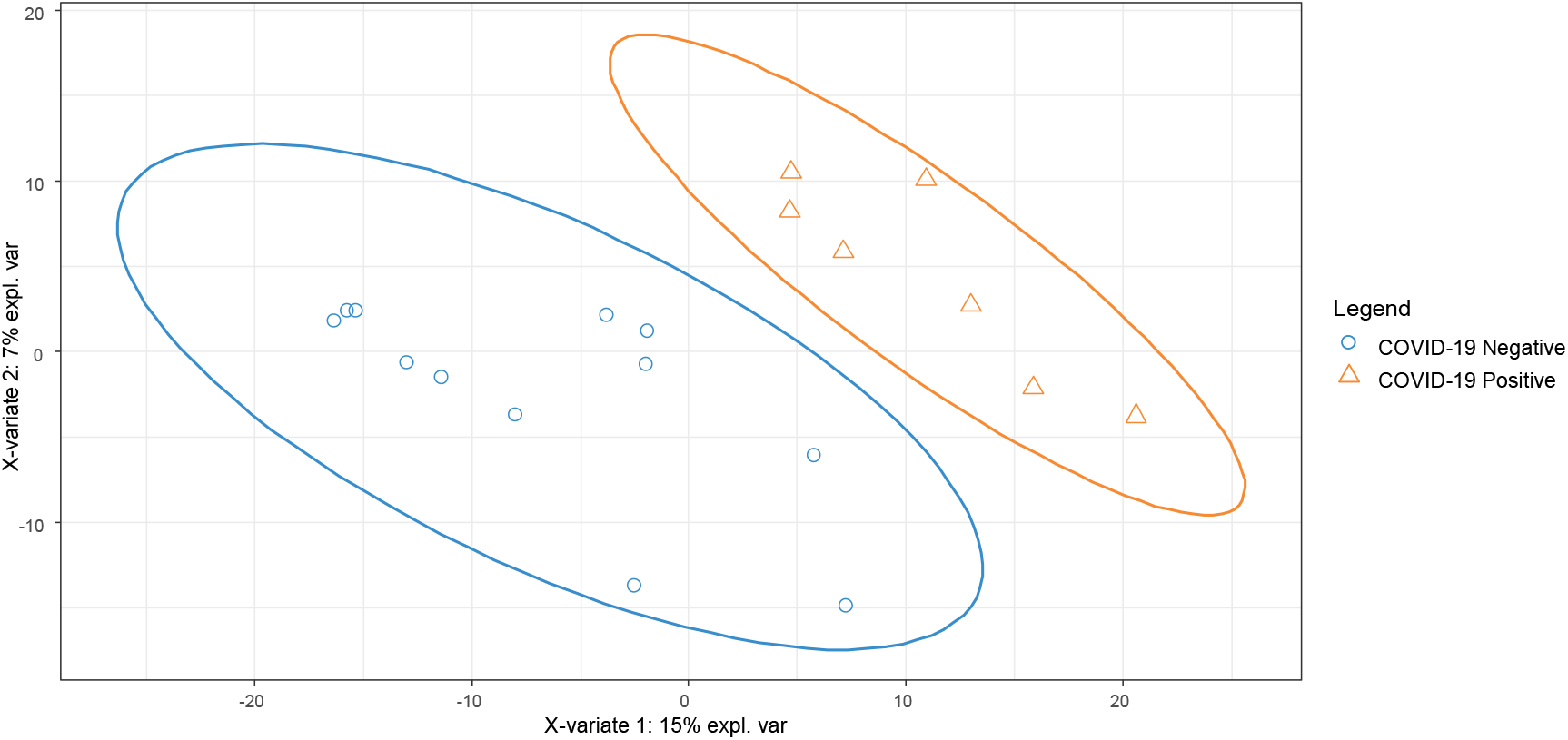
PLS-DA plot for 19 participants treated for T2DM, by COVID-19 positive / negative

**Table S5:** Confusion matrix for COVID-19 positive versus negative (participants taking statins)

| Participants taking statins (n=32) | True COVID-19 Positive (n=11) | True COVID-19 Negative (n=21) |
| --- | --- | --- |
| Predicted COVID-19 Positive (% , n) | 55% (6) | 10% (2) |
| Predicted COVID-19 Negative (% , n) | 45% (5) | 90% (19) |

**Figure S4:**
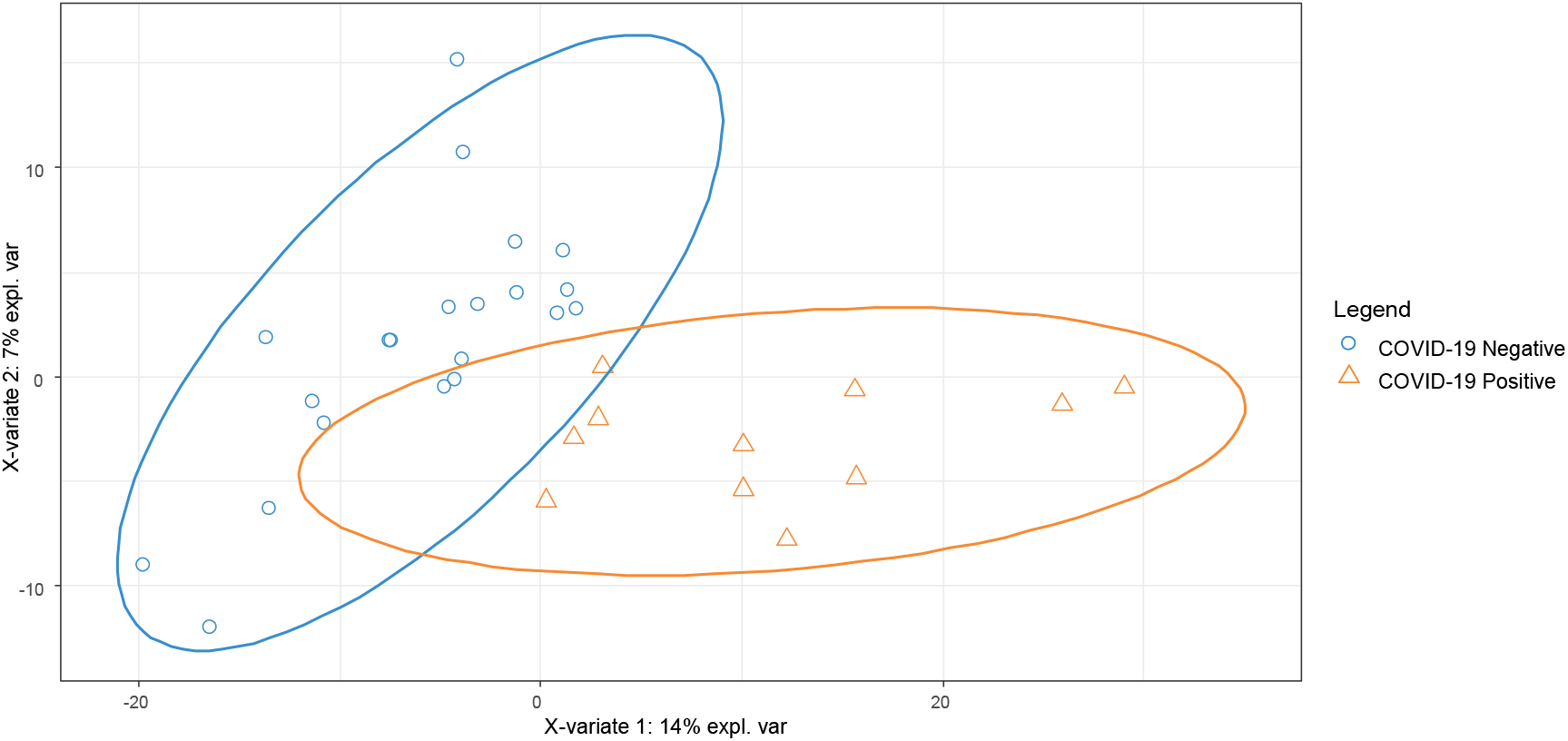
PLS-DA plot for 15 participants treated with statins, by COVID-19 positive / negative

